# Detection of Noroviruses GII in wastewater samples of Bhopal, India, using Droplet Digital PCR

**DOI:** 10.1101/2023.12.13.23299940

**Authors:** R K Nema, J Nagar, A K Singh, A Tandekar, S Singh, A Rahman, O P Sharma, R R Tiwari, D K Sarma, P K Mishra

**Affiliations:** Division of Environmental Biotechnology, Genetics, and Molecular Biology, ICMR- National Institute for Research in Environmental Health, Bhopal - 462030 (MP), India; Division of Environmental Monitoring and Exposure Assessment (Water & Soil), ICMR – National Institute for Research in Environmental Health, Bhopal – 462 030 (MP), India; ICMR – National Institute for Research in Environmental Health, Bhopal – 462 030 (MP), India

**Keywords:** Noroviruses, wastewater-based epidemiology, molecular surveillance, Droplet Digital PCR (ddPCR)

## Abstract

Noroviruses are a significant cause of global gastroenteritis outbreaks, underscoring the importance of effective surveillance. Wastewater-based epidemiology helps identify viral pathogens in communities. Recent advancements in wastewater-based molecular surveillance have shown that viruses can be detected in feces and urine early, making sewage monitoring an essential tool for tracking viral presence. We aimed to create and validate a new method for detecting and monitoring Noroviruses GII in Bhopal’s wastewater using Automated Droplet Digital PCR (ddPCR) technology. In this study, a ddPCR assay targeting the ORF1-2 region of Norovirus GII was developed, allowing viral nucleic acid quantification without a standard curve. A total of 27 samples from five Sewage Treatment Plants located in Bhopal city were collected during the summer season (April and May 2023) at fortnightly intervals and analyzed for the presence of Norovirus using the novel ddPCR assay. Among the samples tested, 33% tested positive for Norovirus, with the highest detection rate observed as 72.72%, followed by 25%. The concentrations of Noroviruses GII in positive samples ranged from 0.06 to 6.60 copies/µl. These findings indicate a potentially higher patient population within Bansal Hospital’s catchment area than the other STPs in the Bhopal region. The study underscores Norovirus’s varying prevalence and distribution in wastewater across different STPs in Bhopal. Moreover, it demonstrates the utility of wastewater surveillance and digital PCR in accurately and specifically detecting Norovirus in wastewater. The practical application of this wastewater surveillance strategy could serve as an early warning system for communities, enabling timely preparedness for impending viral outbreaks, implementation of effective administrative containment measures, and intensified vaccination campaigns.

## Introduction

Norovirus is responsible for around 20% of all diarrhea cases globally, affecting children and adults equally. It is estimated to cause over 200,000 deaths annually in developing countries (1). These highly contagious and widespread RNA viruses are responsible for a notable proportion of global cases of acute gastroenteritis. viral gastroenteritis cases worldwide. The major cause of acute viral gastroenteritis worldwide is rotavirus, with norovirus ranking second in prevalence (2). Noroviruses can cause severe gastroenteritis, with symptoms including nausea, vomiting, diarrhea, and stomach cramps. Noroviruse infection can result in dehydration, especially in young children, older adults, and individuals with compromised immune systems. (3, 4). Noroviruses have the ability to infect not only humans but also a diverse range of other mammalian hosts. For the first time, two previously unidentified norovirus genomes were discovered in raccoon dogs (5). The viruses can survive in the environment for extended periods and are resistant to many common disinfectants, making them difficult to control (3). Infection control measures, such as improved sanitation and hand hygiene, are essential for preventing the spread of noroviruses. One of the ways in which noroviruses can spread is through the fecal-oral route, leading to the contamination of water sources and wastewater (6).

Noroviruses belong to the family Caliciviridae; considering the complete VP1 amino acid sequences, Noroviruses can be genetically classified into ten genogroups labelled as GI to GX. Genogroups I, II, IV, VIII and IX are known to infect humans, with genogroup II (GII) being the most prevalent. Currently, within these ten genogroups, there are 49 confirmed capsid genotypes based on the amino acids of the complete VP1 sequence. These include nine genotypes for GI, 27 for GII, 3 for GIII, 2 for GIV, 2 for GV, 2 for GVI, and one genotype each for GVII, GVIII, GLX, and GX. Moreover, there are 60 confirmed P-types based on partial nucleotide sequences of the RdRp regions, with 14 for GI, 37 for GII, 2 for GIII, 1 for GIV, 2 for GV, 2 for GVI, 1 for GVII, and 1 for GX (7, 8). The GII strain of Norovirus encompasses both porcine and human strains, while GIII consists solely of bovine strains, and GV exclusively contains murine strains (7). Diagnosing a norovirus infection can be challenging due to the presenceexistence of non-specific symptoms and the challenges in diagnosis. However, laboratory testing, particularly molecular assays, has proven to be the most reliable method for diagnosing norovirus infection (9). In addition to laboratory testing, clinical evaluation and epidemiological investigation play crucial roles in diagnosing norovirus infection, especially during outbreaks. These tools help in identifying patterns and sources of infection. Timely diagnosis and the enforcement of control measures are crucial to prevent the further spread of the viruses(10).

Traditional methods for detecting microbial pathogens in water and wastewater often rely on cultivating them in artificial media or cell cultures. However, these methods can be time consuming and less sencitive. Emerging technologies like Polymerase Chain Reaction (PCR) offer a promising alternative. PCR can rapidly and precisely detect a wide range of pathogens from a single water sample, addressing the limitations of traditional methods. In recent years, molecular techniques such as PCR and RT-qPCR have been employed to detect noroviruses in wastewater samples. However, it is important to acknowledge that these methods may have limitations in terms of sensitivity, specificity, and the ability to detect low levels of the virus (11). Digital droplet PCR (ddPCR) represents a cutting-edge approach to nucleic acid detection, boasting several advantages over traditional PCR methods. This innovative technique offers heightened sensitivity, reduced variability, and the unique capability to quantify low levels of target nucleic acids. Beyond these advantages, one of the key features of ddPCR is its versatility in detecting multiple targets simultaneously within a single reaction, making it particularly invaluable for the surveillance of various pathogens, including noroviruses, in wastewater (12). The sensitivity and specificity inherent in ddPCR enable the detection of viruses even at markedly lower concentrations, providing a more profound understanding of viral epidemiology. This breakthrough has significantly impacted clinical research and the surveillance of enteric viruses in wastewater. Molecular surveillance plays a crucial role in understanding the evolution and spread of Norovirus, a highly contagious gastrointestinal pathogen (13). By analysing the genetic sequences of various strains, researchers can acquire insights into the epidemiology, evolution, and ecology of this significant human pathogen.. Ultimately, this research will contribute to developing more effective strategies for preventing norovirus infections in the population.

## Methods

### Waste water sample collection

Sewage samples, 2×500 ml, were collected fortnightly from each of the 5 Waste Water Treatment Plants (WWTP) in the Municipal Corporation Area of Bhopal, Madhya Pradesh. These plants were situated in Bansal Hospital, Charimli, Shirin River, Professor Colony, and Jamuniya Cheer area and had a well-defined catchment area. Samples collected from these areas were brought to the lab and processed for RNA isolation immediately or stored at 4°C and were processed within 24 hours to avoid further degradation of viral nucleic acid.

### Virus concentration using Polyethylene Glycol Precipitation Method

Virus concentration was done using modified polyethylene glycol method (14-16). Initially, the sewage samples (50ml) were centrifuged at 4500 x g for 30 minutes, followed by filtering the supernatant using 0.22µm filters. The pH of the supernatant was then adjusted to 7.0-7.5. Subsequently, 10% PEG 8000 (polyethyleneglycol) and NaCl were introduced to the supernatant to achieve a final concentration of 0.3 Molar. The blend was suxequently left to incubater overnight at 4°C followed by centrifugation at 4500x g for 30 minutes. The obtained supernatant was discarded, and the resulting pellets were resuspended in 500µL of phosphate buffer saline solution (PBS) to form a virus concentrate. The virus concentrate can either be processed for nucleic acid extraction or stored at −80°C for long-term storage until extraction.

### RNA extraction

Total RNA was extracted from an aliquot of each viral concentrate by AllPrep Power Viral DNA/RNA Kit (Qiagen, Germany), following the manufacturer’s instructions. This technique selectively bound viral RNA to a silica-gel membrane, while contaminants were efficiently eliminated during washing steps, removing PCR inhibitors such as divalent cations and proteins (17-19). Extracted RNA was eluted in 100 µL of the elution buffer and stored in 10 µL aliquots at-80°C until use.

### cDNA Synthesis

The cDNA was synthesized using the cDNA Synthesis Kit GoScript^™^ Reverse Transcription System (Cat. No. A5000) (Promega), following the modified manufacturer’s protocol (20, 21). Briefly, 4 μl of RNA and 1 μl of 0.2µg/µl random hexamer primer underwent denaturation at 70°C for 5 min. A 15 µl reverse transcription mix, including 5x reaction buffer, Ribonuclease Inhibitor, dNTP mix, and RT enzyme, was added to the RNA/primer mix. After proper mixing and brief centrifugation, the tube underwent incubation at 25°C for 5 min, followed by 42°C for 60 min for cDNA synthesis. Termination of RT reaction was achieved at 70°C for 10 min. The tubes were subsequently chilled on ice, and the cDNA was stored at −80°C for future use.

### ddPCR Assay

Primers and TaqMan probe targeting the highly conserved ORF1, 2 regions of the Norovirus GII genotype were synthesized. The specificity of these selections was validated using the BLAST program from NCBI. Notably, the TaqMan probe was tagged with 5’-FAM for the fluorescent reporter dye and 3’-BHQ-1 as the quencher, ensuring assurate signal detection in the assay.. The sequences of the primers and probes are shown in Table 1. The ddPCR reaction was prepared using 2µl (50 ng) of cDNA and a reaction mixture that included ddPCR Supermix for probes, forward and reverse primers, and the TaqMan probe. Droplet generation was then carried out using an Automatic Droplet Generator, QX200^™^ AutoDG™, in a 96-well polypropylene plate. The subsequent PCR cycling involved initial denaturation, followed by 45 cycles of denaturation (95°C for 15 seconds), annealing, and extension(55°C for 30 seconds), culminating in a final denaturation step (55°C for 7 minutes). After PCR, the plate containing the droplets was transferred to a QX200 droplet reader for analysis. Finally, data analysis was performed using QuantaSoft^™^ Analysis Pro software, which involved discriminating positive from negative droplets based on fluorescence amplitude, thereby identifying the positive droplets containing the amplification products. This comprehensive approach ensured precise quantification and high-resolution analysis of the Norovirus genetic material.

**Table 1:**
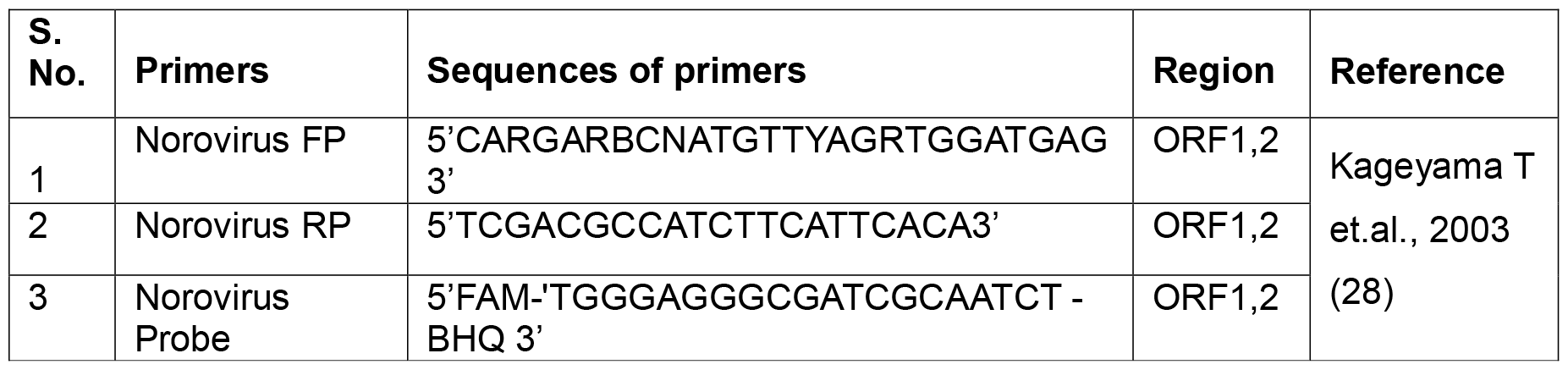
Primers and probe used in the development of ddPCR assay for Noroviruses.

## Result

A ddPCR assay targeting the ORF1-2 region of Norovirus GII wasdeveloped, allowing viral nucleic acid quantification without a standard curve. A total of 27 sewage samples were collected from five Sewage Treatment Plants (STPs) located in Bhopal for the purpose of investigating the presence of Norovirus using the highly sensitive norovirus droplet digital polymerase chain reaction (ddPCR) assay. Among these samples, nine were confirmed to be positive for Norovirus using the ddPCR assay. In particular, within the Bansal Hospital STP subset, consisting of 11 samples, a significant majority of 8 samples (72.72%) exhibited a positive result for Norovirus when subjected to the ddPCR assay. Similarly, within the Char Imli STP subset, encompassing four samples, one sample (25%) was found to be positive for Norovirus through the ddPCR assay (Table 2). The concentrations of Noroviruses GII in positive samples ranged from 0.06 to 6.60 copies/μl (Table 3). However, the samples collected from Professor Colony, Jamunia Cheer, and Sirin River, representing distinct geographic locations, yielded negative results for Norovirus in the ddPCR assay. This suggests the absence of detectable Norovirus within the samples obtained from these areas.

**Table 2:**
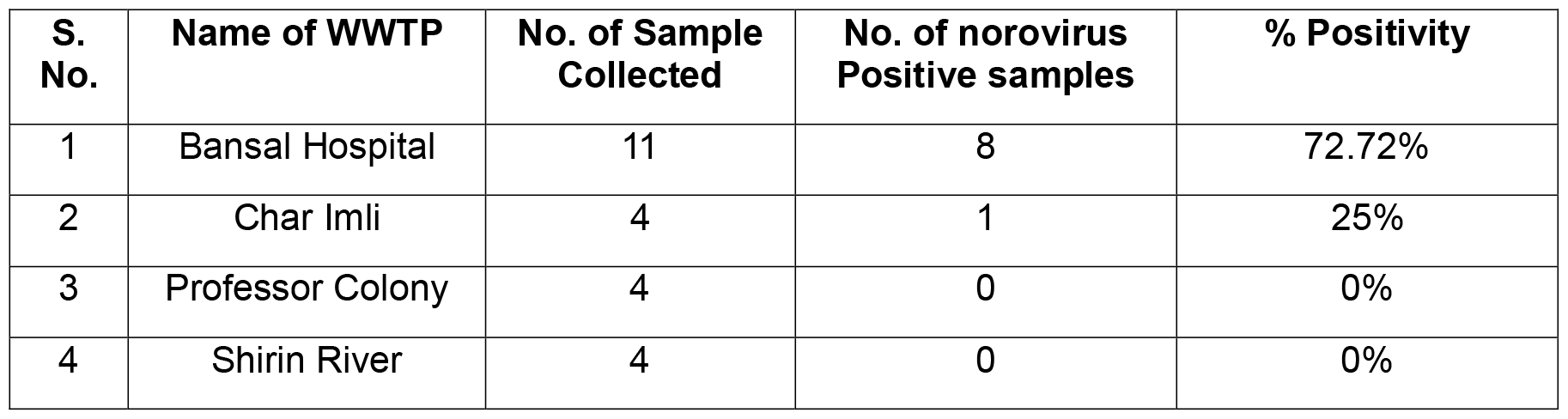
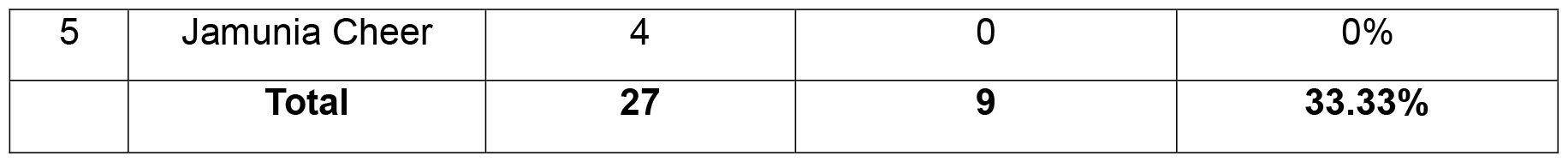
Distribution of Norovirus in wastewater treatment plants of Bhopal City.

**Table 3:** Concentration of Noroviruses GII in positive samples.

| **S. No.** | **Date of sample collection** | **Name of STPs** | **No. of DDPCR Positive Droplets** | **Concentration (Copies/ μl)** |
| --- | --- | --- | --- | --- |
| 1 | 19.04.2023 | Bansal Hospital | 96 | 6.60 |
| 2 | 22.04.2023 | Char Imli | 17 | 0.94 |
| 3 | 22.04.2023 | Bansal Hospital | 58 | 3.53 |
| 4 | 23.04.2023 | Bansal Hospital | 27 | 1.64 |
| 5 | 24.04.2023 | Bansal Hospital | 35 | 2.23 |
| 6 | 25.04.2023 | Bansal Hospital | 36 | 2.44 |
| 7 | 26.04.2023 | Bansal Hospital | 01 | 0.06 |
| 8 | 27.04.2023 | Bansal Hospital | 12 | 0.77 |
| 9 | 28.04.2023 | Bansal Hospital | 03 | 0.20 |

## Discussion

The augmented Norovirus positivity observed in this study is hypothesized to be associated with the presence of a hospitalized patient population. This correlation could plausibly lead to an increased likelihood of Norovirus shedding in feces and urine among the affected individuals(22). As a result, the sewage system may experience an elevation in the viral load, potentially contributing to the heightened Norovirus prevalence in the studied area. Further investigation is warranted to validate and understand the implications of these findings on public health and environmental management.

The results of this study offer valuable insights into the prevalence of Norovirus in Sewage Treatment Plants (STPs) located in Bhopal, India. The detection and analysis of Norovirus using the droplet digital polymerase chain reaction (ddPCR) assay yielded significant results, highlighting the importance of understanding the presence and distribution of this pathogen in wastewater (23).

The overall norovirus detection rate of approximately 35.7% among the collected samples indicates a significant presence of Norovirus in the sewage samples analyzed. These findings are consistent with prior research that has recognized wastewater as a potential reservoir for the transmission of norovirus (24). The high detection rate emphasizes the importance of wastewater surveillance for monitoring and managing potential health risks associated with norovirus contamination.

The Bansal Hospital STP demonstrated the highest norovirus detection rate, with 72% of the samples testing positive for the virus. This finding suggests a higher prevalence and potential concentration of Norovirus in the sewage originating from this specific treatment plant. The increased detection rate in this STP might be attributed to factors such as a higher number of infected patients admitted to Bansal Hospital, contributing to the viral load in the wastewater treatment plants. Further investigation is necessary to identify the specific factors influencing the higher detection rate in this particular STP. Conversely, the Char Imli STP exhibited a lower norovirus detection rate of approximately 25%. Although the overall detection rate is lower than the Bansal Hospital STP, it still signifies Norovirus’s presence in the analyzed wastewater samples. The differences observed in detection rates between the two STPs may be attributed to variations in the sources of wastewater, treatment processes, or geographical factors influencing the viral load in the sewage. Understanding these variations is crucial for implementing effective control measures and optimizing treatment processes to minimize norovirus transmission. Interestingly, no norovirus was detected in the samples collected from Professor Colony, Jamunia Cheer, and Sirin River. This result suggests either the absence of norovirus contamination in these specific locations or the presence of low viral loads that fell below the detection threshold of the ddPCR assay. Further investigations using alternative detection methods, such as quantitative PCR or sequencing, could provide a more comprehensive understanding of norovirus presence in these areas (25).

The findings of this study underscore the importance of robust surveillance programs and the implementation of adequate wastewater treatment processes to mitigate norovirus transmission. Wastewater-based epidemiology, including the monitoring of sewage for the presence of pathogens like Norovirus, can serve as an early warning system for outbreaks and provide valuable information for public health interventions. Improvements in wastewater treatment processes, particularly in STPs with higher detection rates, should be considered to ensure the effective removal of Norovirus and minimize its potential spread through environmental contamination.

Furthermore, the identification of norovirus-positive samples in STPs highlights the importance of public awareness and hygiene practices to reduce the risk of infection. Proper hand hygiene, food handling, and the disinfection of contaminated surfaces are crucial in preventing norovirus outbreaks in communities(26). To strengthen the validity and applicability of research findings, future efforts should prioritize recruiting a larger and more diverse sample.. Additionally, sequencing and phylogenetic analysis of the norovirus strains detected in the wastewater samples would provide insights into the genetic diversity and potential sources of the virus.

## Data Availability

All data produced in the present work are contained in the manuscript

